# Fuzzy Recurrence Dynamics of Contrast Medium Extravasation in Computed Tomography

**DOI:** 10.1101/2025.03.10.25323651

**Authors:** Tuan D. Pham, Maki Kitamura, Taichiro Tsunoyama

## Abstract

**Objectives:** The discovery of complex patterns in contrast medium extravasation on computed tomography (CT) imaging is critical for improving trauma patient management. Identifying these patterns enables early detection of complications such as vascular injury, organ rupture, and active hemorrhage, facilitating timely and targeted interventions that enhance patient outcomes. This study introduces an advanced imaging analytics approach that integrates nonlinear dynamic analysis and geostatistical methods to characterize the temporal and spatial evolution of contrast medium extravasation in trauma cases.

**Methods:** We analyzed CT imaging sequences from trauma patients using fuzzy recurrence dynamics to uncover hidden structures within contrast dispersion patterns. This methodology quantifies subtle variations in blood flow, capturing previously unrecognized radiographic signatures associated with hemodynamics. Recurrence-based metrics were leveraged to identify dynamic changes indicative of impending complications, enhancing the predictive capabilities of trauma imaging.

**Results:** The proposed approach effectively detected subtle, high-risk extravasation patterns that are often overlooked by conventional imaging techniques. The integration of nonlinear dynamic analysis and geostatistical modeling provided a more precise characterization of contrast dispersion, revealing predictive markers of vascular compromise. These findings support the application of advanced computational techniques for improving trauma imaging and clinical decision-making.

**Conclusion:** The findings demonstrate the potential of integrating advanced nonlinear dynamics and network techniques into trauma imaging, offering a new framework for real-time detection, risk stratification, and predictive modeling of extravasation events. This approach represents a step toward precision medicine in emergency care, enabling automated, data-driven decision support for clinicians. By improving diagnostic accuracy and facilitating early therapeutic intervention, this study lays the foundation for a paradigm shift in trauma imaging, ultimately optimizing patient management and outcomes in critical care settings.

## 1 Introduction

Contrast media extravasation [1] occurs when intravenously administered contrast agents leak from the vascular system into surrounding soft tissues, often due to vessel injury or increased vascular permeability. The severity of this complication varies, ranging from mild discomfort and localized swelling to severe outcomes such as compartment syndrome, tissue necrosis, and long-term functional impairment [2]. Key factors influencing tissue damage include the volume and type of contrast agent, injection pressure, and the patient’s underlying health conditions. Despite being one of the most common adverse events in radiology, contrast extravasation remains understudied, particularly regarding its dynamic progression and predictive markers [1, 2].

The ability to detect contrast medium extravasation in computed tomography (CT) imaging is crucial in trauma patients, as early identification facilitates timely surgical or interventional management, reducing morbidity and mortality [4, 5]. Advanced CT techniques, including multi-phase imaging and contrast-enhanced scans [6, 7], improve visualization, allowing clinicians to assess extravasation extent and localize vascular injuries with greater accuracy. Additionally, differentiating contrast extravasation from hemorrhage is essential, as it directly influences treatment decisions. The evolving literature underscores the importance of precise imaging protocols and computational tools for enhancing diagnostic accuracy and clinical decision-making.

Contrast extravasation is often identified by hyperdense areas on CT scans. For instance, Mericle et al. [8] defined contrast extravasation as a hyperdensity with a maximal Hounsfield unit measurement exceeding 90 and/or the disappearance of the hyperdensity on a repeat CT within 24 hours. This variability in definitions necessitates a standardized approach to enhance diagnostic accuracy [9]. The detection of extravasation is particularly challenging in specific surgical interventions, such as kyphoplasty. Palm et al. [10] noted that even minimal extravasation can have significant clinical implications, yet traditional imaging techniques like fluoroscopy may fail to detect small leakages, necessitating the use of advanced modalities such as intraoperative CT.

CT also plays a crucial role in assessing anastomotic leaks post-surgery. Studies have shown that mediastinal air or fluid collections can serve as indicators of anastomotic leakage, with sensitivity varying significantly. Plat et al. [11] demonstrated that air and fluid collections in the mediastinum had predictive value for assessing anastomotic leakage. Additionally, Kim et al. [12] found that intraluminal contrast-enhanced CT significantly improves the detection of gastrointestinal leaks, achieving a diagnostic performance of 96.6%. This highlights the critical role of contrast-enhanced CT in postoperative assessments, particularly for gastrointestinal tract complications. Differentiating contrast extravasation from hemorrhage is another essential aspect of CT imaging. Yedavalli and Sammet [13] discussed the diagnostic challenges posed by hyperdensities on post-procedural imaging in stroke patients, which may indicate either extravasation or intracranial hemorrhage. This distinction is vital, as it influences clinical management strategies. The authors advocated for improved imaging protocols to reduce the risk of misdiagnosis and inappropriate treatment.

Thus, the detection of contrast medium extravasation in CT is a critical aspect of radiological practice, particularly in trauma assessments and various surgical procedures. Chronological literature highlights evolving methodologies and diagnostic criteria, underscoring the need for accurate imaging techniques to differentiate extravasation from other complications [14, 15, 16, 17, 18, 19, 20, 21, 22, 23, 24].

A deeper understanding of extravasation dynamics can provide novel insights into trauma-related injuries, facilitate early complication prediction, and support personalized diagnostic and interventional strategies. Identifying complex extravasation patterns in CT imaging could enable real-time detection of high-risk cases, improving early intervention and optimizing patient outcomes. Computational approaches incorporating geostatistics [25], nonlinear dynamics and recurrence-based analysis [26, 27] can offer promising avenues for uncovering hidden structures within contrast dispersion patterns.

Building on recent research [28] employing geostatistics, artificial intelligence-based non-linear dynamics, and recurrence network analysis to identify gender-specific radiographic features in CT scans, this study extends prior work [29] by investigating nonlinear dynamic features in CT imaging of extravasation in trauma cases. By analyzing the spatial and temporal evolution of extravasation, the study aims to uncover radiographic patterns that serve as predictive markers for clinical outcomes. Integrating geostatistical modeling and fuzzy recurrence networks enables a detailed characterization of contrast dispersion, revealing subtle anomalies indicative of complications such as hemorrhage or vascular compromise. These insights could enhance diagnostic precision, facilitate early therapeutic interventions, and improve the accuracy and timeliness of trauma care. Furthermore, this research contributes to developing advanced computational frameworks that integrate artificial intelligence for automated pattern recognition, paving the way for more effective and personalized clinical decision-making in emergency radiology.

The remainder of this paper is structured as follows: Section 2 details the mathematical foundations of semivariogram modeling, fuzzy recurrence analysis, and fuzzy recurrence networks used in this study. Section 3 presents the findings, highlighting the nonlinear dynamic patterns of extravasation observed in three trauma cases. Section 4 provides a comprehensive discussion of the implications, and Section 5 summarizes the conclusions and potential impact of the results.

## 2 Methods

### 2.1 The semivariogram function

In geostatistics [25, 30, 31, 32], the semivariogram function quantifies the spatial dependence of a regionalized variable. Given a spatial domain where measurements of a random field *z*(*i*) are taken at different locations *i*, the semivariogram function is defined as:

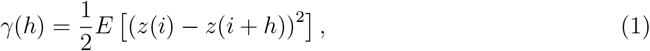

where *h* is the separation distance (lag) between two spatial locations, *E*[·] denotes the expectation operator, and *z*(*i*) and *z*(*i* + *h*) represent values of the spatial variable at locations *i* and *i* + *h*, respectively.

The semivariogram is used in this study to assess the spatial variability of intensity values within a CT slice exhibiting extravasation, where *z*(*i*) = *I*(*x, y*) represents the intensity of a pixel at coordinates (*x, y*) in the 2D CT image.

For empirical estimation, the semivariogram is computed from a set of spatial observations as:

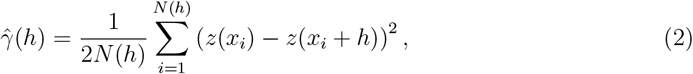

where *N* (*h*) is the number of pairs of observations separated by the distance *h*. A common theoretical model for the semivariogram is the spherical model:

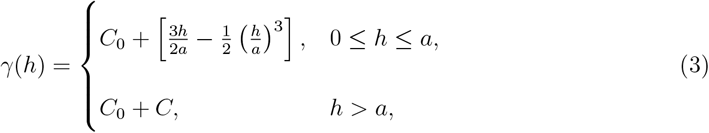

where *C*_0_ is the nugget effect, *C* is the sill (the total variance), and *a* is the range, beyond which spatial correlation becomes negligible. The nugget effect refers to the discontinuity at the origin of the semivariogram function. It represents the small-scale variability or measurement error that cannot be captured by the spatial model. Mathematically, if the semivariogram *γ*(*h*) does not approach zero as the lag distance *h* → 0, the semivariogram exhibits a nugget effect.

### 2.2 Fuzzy recurrence plots

A traditional recurrence plot (RP), denoted as **R** is used to analyze the recurrence behavior of a time series by detecting when states in a reconstructed phase space are sufficiently close. It is defined as:

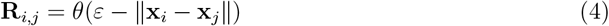

where **x**_*i*_, **x**_*j*_ are state vectors reconstructed from the time series, *ε* is a predefined threshold, *θ*(·) is the Heaviside step function, which assigns a value of 1 if the distance is within the threshold and 0 otherwise.

The fuzzy recurrence plot (FRP) [26] extends the RP by incorporating fuzzy set theory to represent the degree of recurrence rather than a binary assignment. This approach enhances the sensitivity of recurrence analysis, making it more robust to capturing subtle patterns in dynamical systems.

Let **z** = (*z*_1_, *z*_2_, … , *z*_*T*_ ) represent the vectorized pixel intensities of a CT slice, where each element *z*_*i*_ corresponds to the intensity value of a pixel in a one-dimensional representation of the image. To analyze the temporal and spatial structures within the image, we construct its phase space using time-delay embedding.

The phase space representation of the pixel intensity sequence is constructed using an embedding dimension *m* and a time delay *τ* . The resulting phase space, denoted as *mathbf S*, is given by:

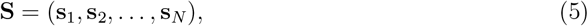

where the number of constructed state vectors is

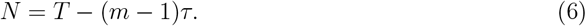

Each state vector **s**_*i*_ is defined as:

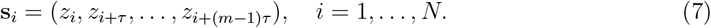

This phase space representation allows the analysis of the underlying dynamical structure in the image by considering the evolution of pixel intensities in a higher-dimensional space.

The FRP, denoted as **F**, quantifies the degree of recurrence between state vectors in the phase space and is constructed by applying the fuzzy *c*-means (FCM) clustering algorithm [33] and fuzzy relations [34] to the phase space representation **S**. It is mathematically defined as

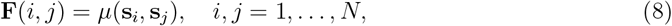

where *µ*(**s**_*i*_, **s**_*j*_) represents the degree of similarity between the state vectors **s**_*i*_ and **s**_*j*_, which is determined using the following three properties:

1. *Self-similarity:*

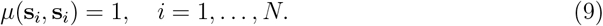
2. *FCM-induced symmetry:*

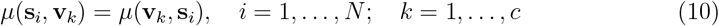

where 0 *< µ*(**s**_*i*_, **v**_*k*_) *<* 1 represents the real membership value, indicating the degree to which **s**_*i*_ belongs to cluster **v**_*k*_. This membership value is computed using the FCM, as described in [26].
3. *Fuzzy relation-based transitivity:*

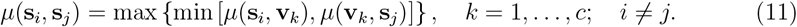

By leveraging fuzzy recurrence analysis, the FRP provides a nuanced characterization of spatial and temporal variations in contrast dispersion. This enables the identification of subtle structures in contrast medium extravasation patterns within CT images. The graded recurrence values help differentiate normal dispersion from pathological changes, contributing to more accurate diagnostics and predictive modeling.

### 2.3 Fuzzy recurrence quantification

Building upon the principles of recurrence plot quantification [36, 37, 38, 39], fuzzy recurrence quantification analysis [35] extends these concepts by introducing a set of fuzzy recurrence metrics, including fuzzy recurrence rate (fRR), fuzzy determinism (fDET), fuzzy laminarity (fLAM), fuzzy trapping time (fTT), fuzzy divergence (fDIV), and fuzzy recurrence entropy (fENT).

An FRP **F** can be transformed into a binary FRP **B** using an image segmentation method described in [35]. The information contained in both **F** and **B** can be utilized to quantify the fuzzy recurrence properties of a dynamical system, as outlined below.

The fRR quantifies the overall density of recurrence points in the FRP and is given by

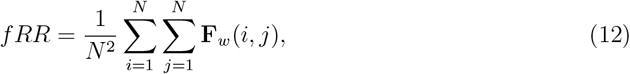

where

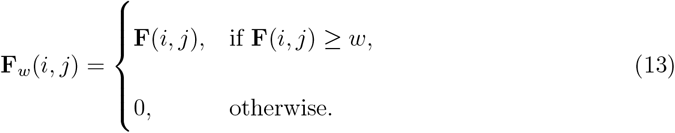

where, *w* represents the threshold for recurrence point selection.

The fDET measures the proportion of recurrence points forming diagonal structures, reflecting the predictability of system dynamics:

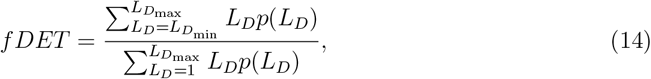

where *L*_*D*_ denotes the diagonal line length, 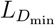 is the minimum diagonal length, and 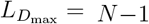 represents the maximum diagonal length of **B**. The function *p*(*L*_*D*_) denotes the probability distribution of diagonal line lengths, obtained from a histogram with 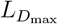 bins.

The fLAM captures the proportion of recurrence points forming vertical structures, indicative of laminar (intermittent) states:

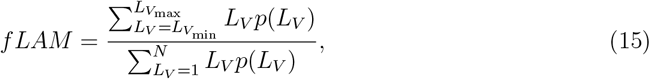

where *L*_*V*_ denotes the vertical line length, 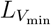 is the minimum vertical length, and 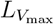 is the maximum vertical length in **B**. The function *p*(*L*_*V*_ ) represents the probability distribution of vertical line lengths, obtained from a histogram with 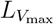 bins.

The fTT represents the average length of vertical line segments in the FRP, providing insight into the duration of laminar states:

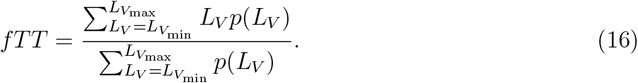

The fDIV is defined as the inverse of the longest diagonal line length, serving as an indicator of system divergence and instability:

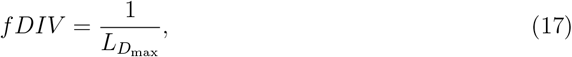

where 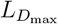 is the maximum diagonal length of **B**.

Finally, the fENT characterizes the complexity of the system by computing the Shannon entropy of the distribution of diagonal line lengths in **B**, providing a measure of dynamical variability:

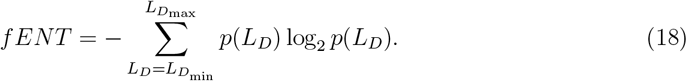

### 2.4 Fuzzy recurrence networks

In this context, the concept of fuzzy recurrence networks (FRNs) [40] is employed to construct network topology and compute graph properties of the extravasation on CT imaging. Utilizing the same fuzzy similarity relations [34], the membership grades of recurrence or similarity between cluster pairs (**v**_*k*_, **v**_*q*_) are defined as follows:

1. *Self-similarity:*

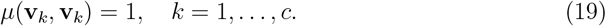
2. *Symmetry:*

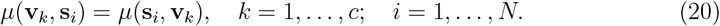
3. *Transitivity:*

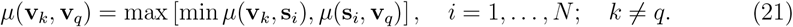

A defuzzification procedure is applied to transform an FRN into a binary network using the *β*-cut method:

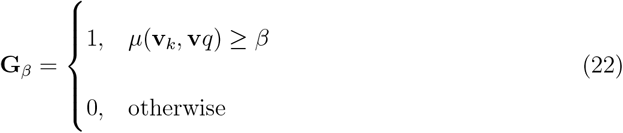

where **G**_*β*_ is a *c* × *c* binarized matrix, and *β* ∈ [0, 1] is the chosen threshold.

Finally, the adjacency matrix of an unweighted *β*-cut recurrence network, denoted as **A**_*β*_, is defined as

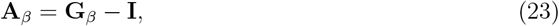

where **I** is the identity matrix, and **A**_*β*_ can be used to compute the characteristic path length and average clustering coefficient of the graph [41, 42].

### 2.5 The largest recurrence eigenvalue

The largest recurrence eigenvalue [43], denoted as *λ*_max_, is determined through a structured process involving convolutional transformations and pooling operations. The procedure is outlined as follows:

1. Initialize the recurrence matrix **F** of size *N* × *N* .
2. Define the filter kernel, activation function (ReLU), pooling size, and stride.
3. Set the final target size as an *n* × *n* convolved matrix, denoted as *c***F**.
4. Iterate while *N > n*:
  a. Apply convolution on **F** using the ReLU activation function.
  b. Perform max pooling on the resulting convolved matrix *c***F**.
5. Terminate the loop when *N* ≤ *n*.
6. Compute *λ*_max_ from the final *c***F**.

The rectified linear unit (ReLU), denoted as *r*(*u*), is applied element-wise and defined as:

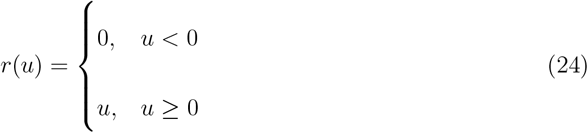

Max pooling is performed to reduce dimensionality while preserving dominant features. Given a pooling size of *δ* × *δ*, we define a set of pooling regions as

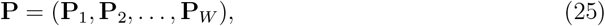

where each pooling region in **P** is represented as

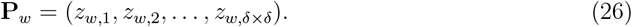

The max pooling function *H*_max_ operates on each pooling region as follows:

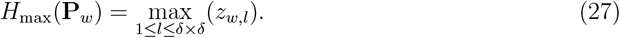

The total number of pooling regions in the convolved matrix *c***F** is determined by the chosen pooling size and stride, which dictate the step size for traversing *c***F**.

## 3 Results

Figures 1, 2, 3, and 4 illustrate representative CT slices demonstrating contrast extravasation in four different trauma cases: liver injury, retroperitoneal hematoma (RPH), splenic trauma, and facial trauma, respectively. The regions of interest containing extravasations were manually extracted and subsequently resized to 30 × 30 pixels. This upscaling, while preserving the structural integrity of the extravasation patterns, was performed to ensure compatibility with downstream computational analyses, including geostatistical and recurrence-based quantification.

**Figure 1:**
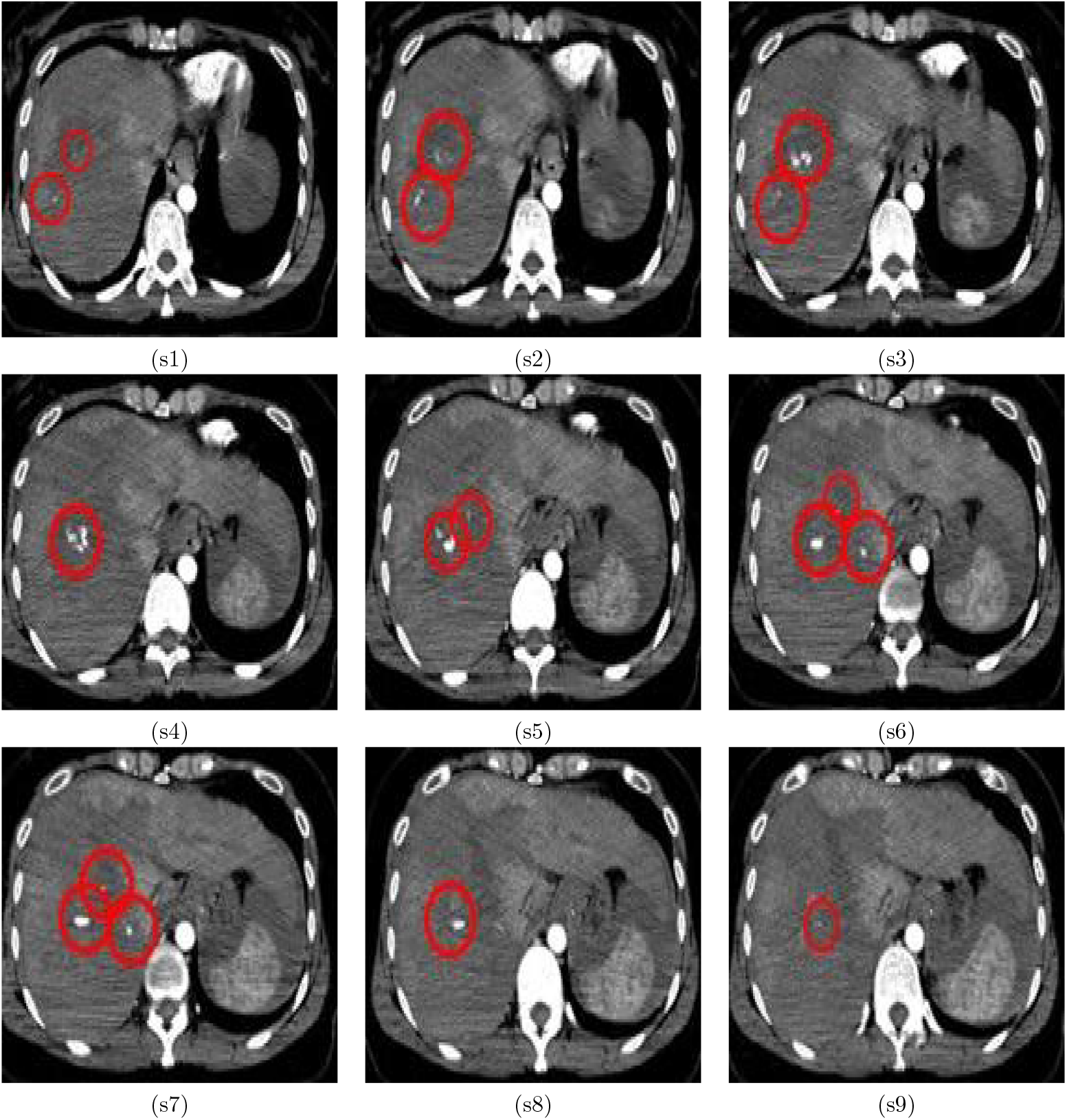
Sequential CT showing extravasation (marked with circles) within the liver, where s1, …, s9 indicate slice #1, …, slice #9.

**Figure 2:**
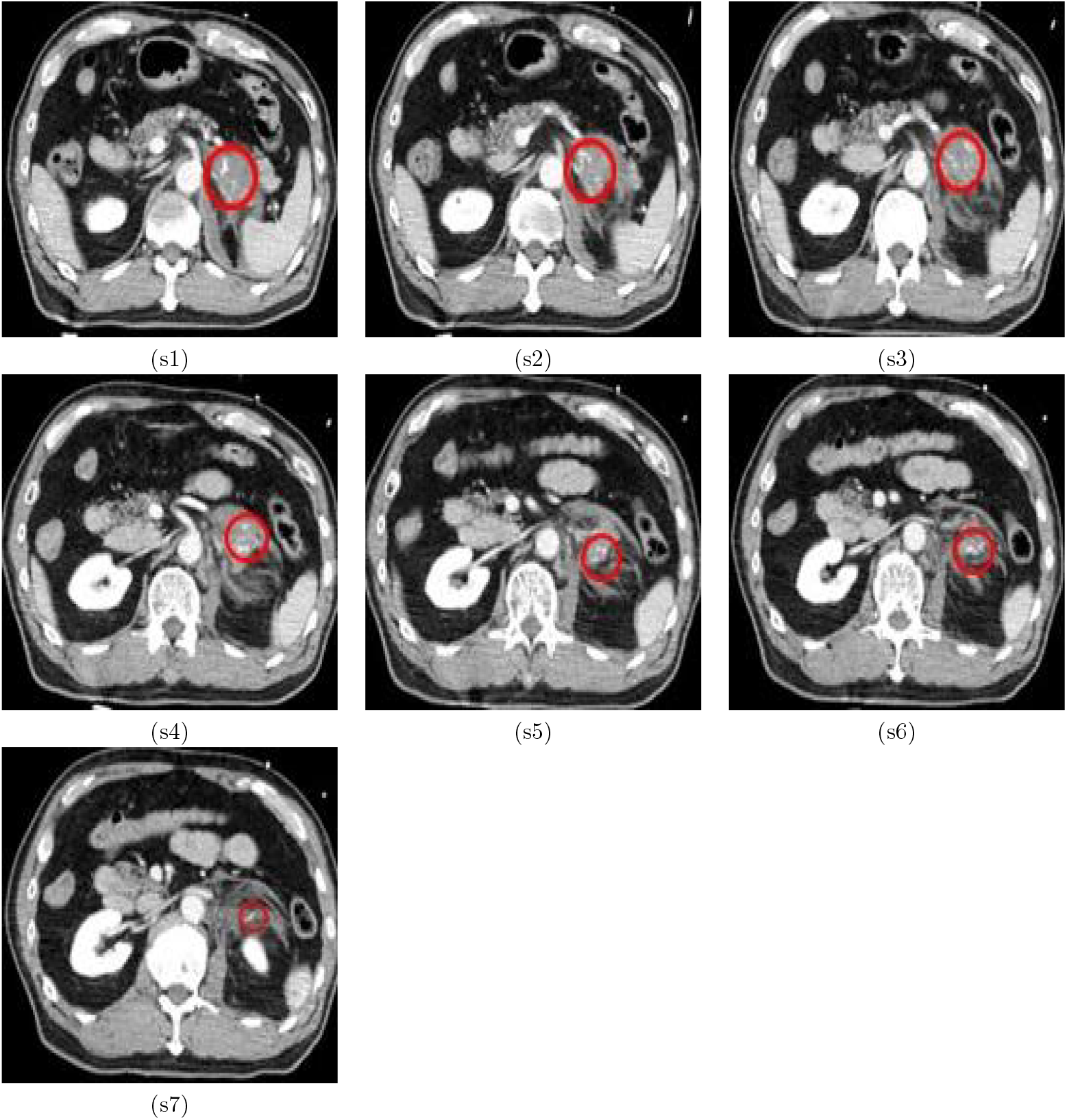
Sequential CT showing extravasation (marked with circles) in a retroperitoneal hematoma, where s1, …, s7 indicate slice #1, …, slice #7.

**Figure 3:**
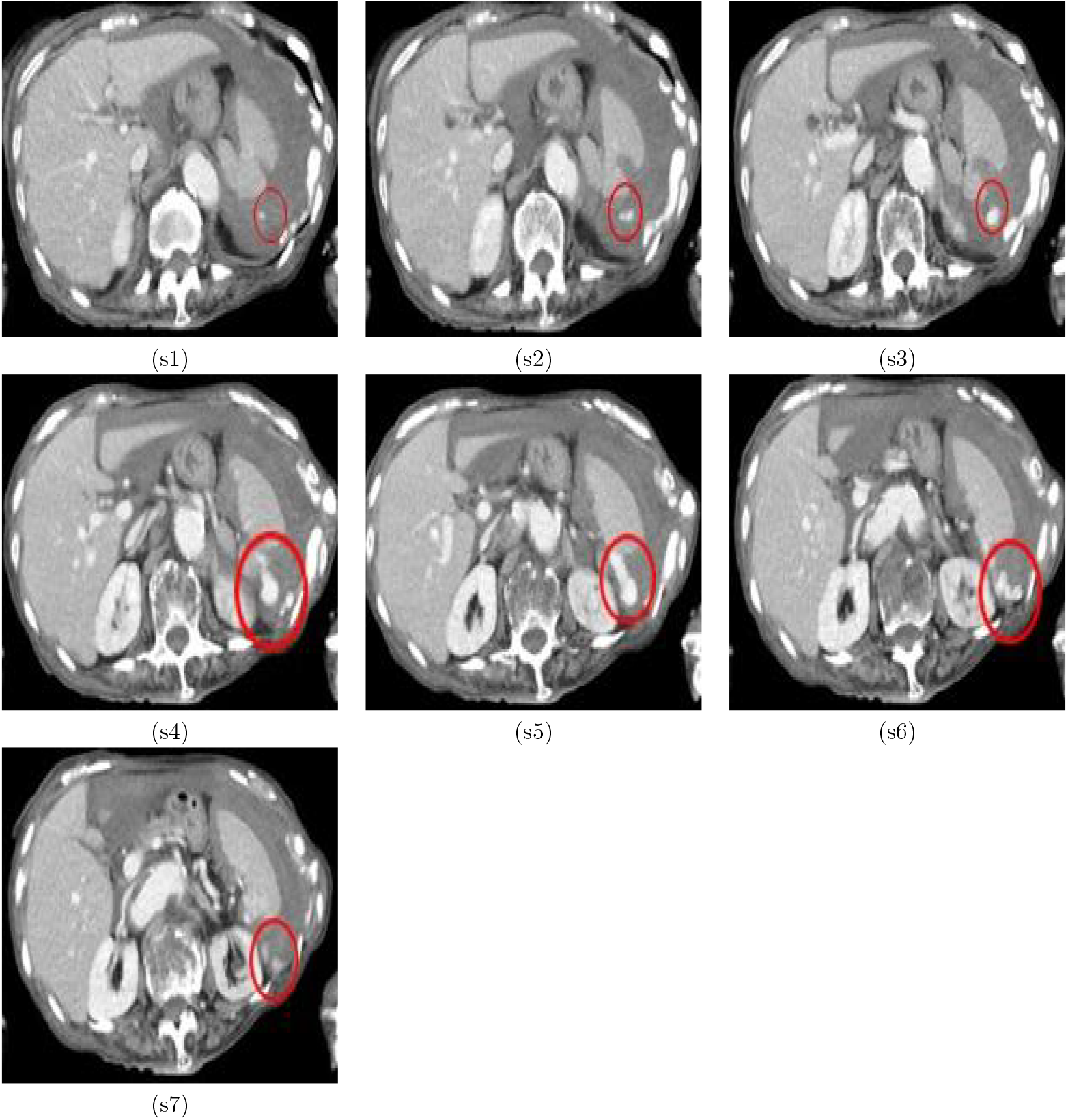
Sequential CT showing extravasation (marked with circles) within the spleen, where s1, …, s7 indicate slice #1, …, slice #7.

**Figure 4:**
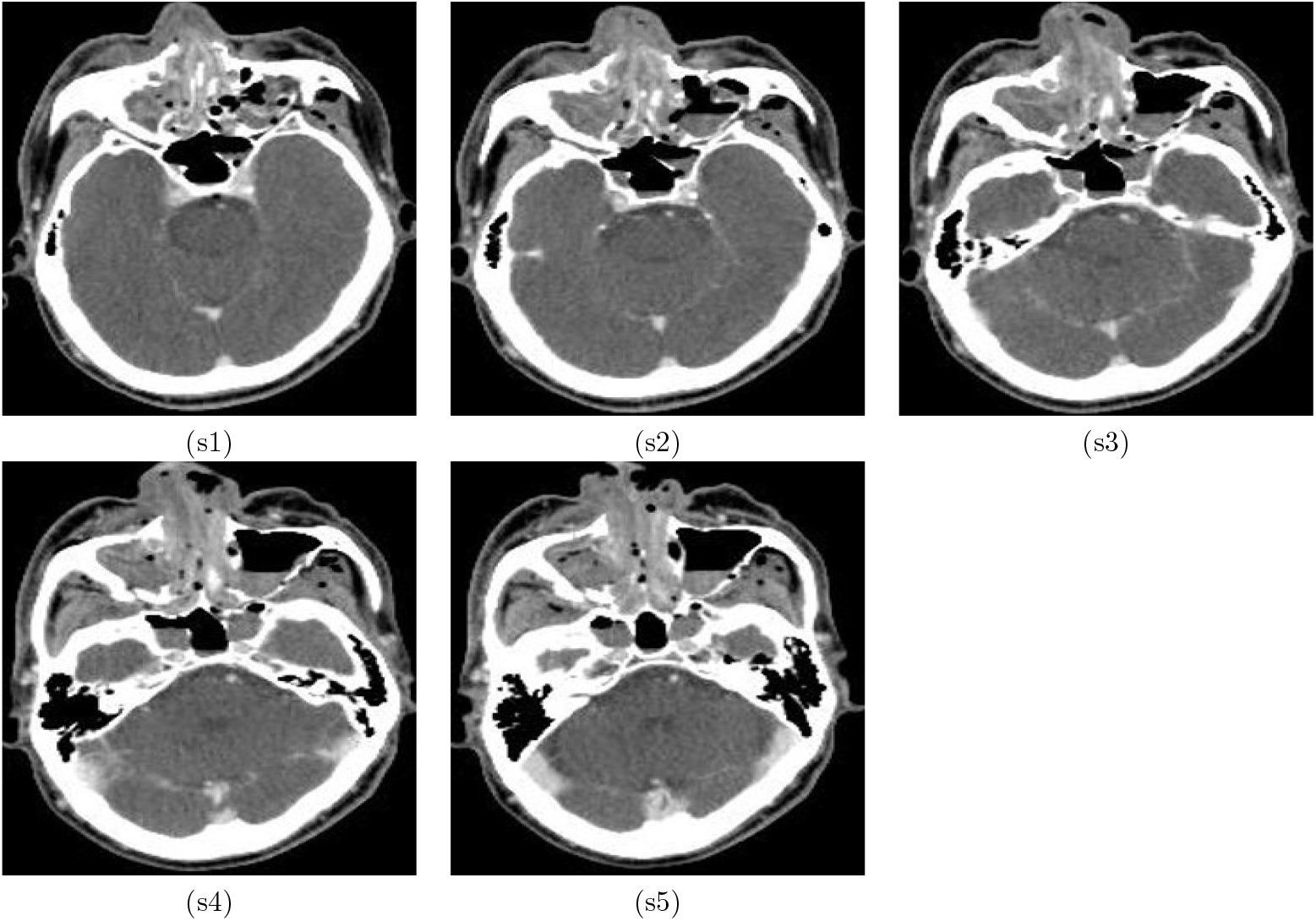
Sequential CT showing extravasation (marked with circles) within the facial trauma, where s1, …, s5 indicate slice #1, …, slice #5.

To assess the spatial heterogeneity of extravasation patterns, Figure 5 presents the empirical semivariograms computed for each trauma case. The semivariograms capture the spatial correlation of intensity variations within the extravasation regions, providing insight into local tissue disruption and contrast dispersion. Empirical semivariograms were preferred over theoretical models (e.g., spherical) as they could directly capture the spatial variance of contrast extravasation from observed CT data without assuming predefined structures. This is crucial for modeling heterogeneous and anisotropic patterns in medical imaging, where theoretical models may oversimplify spatial correlations.

**Figure 5:**
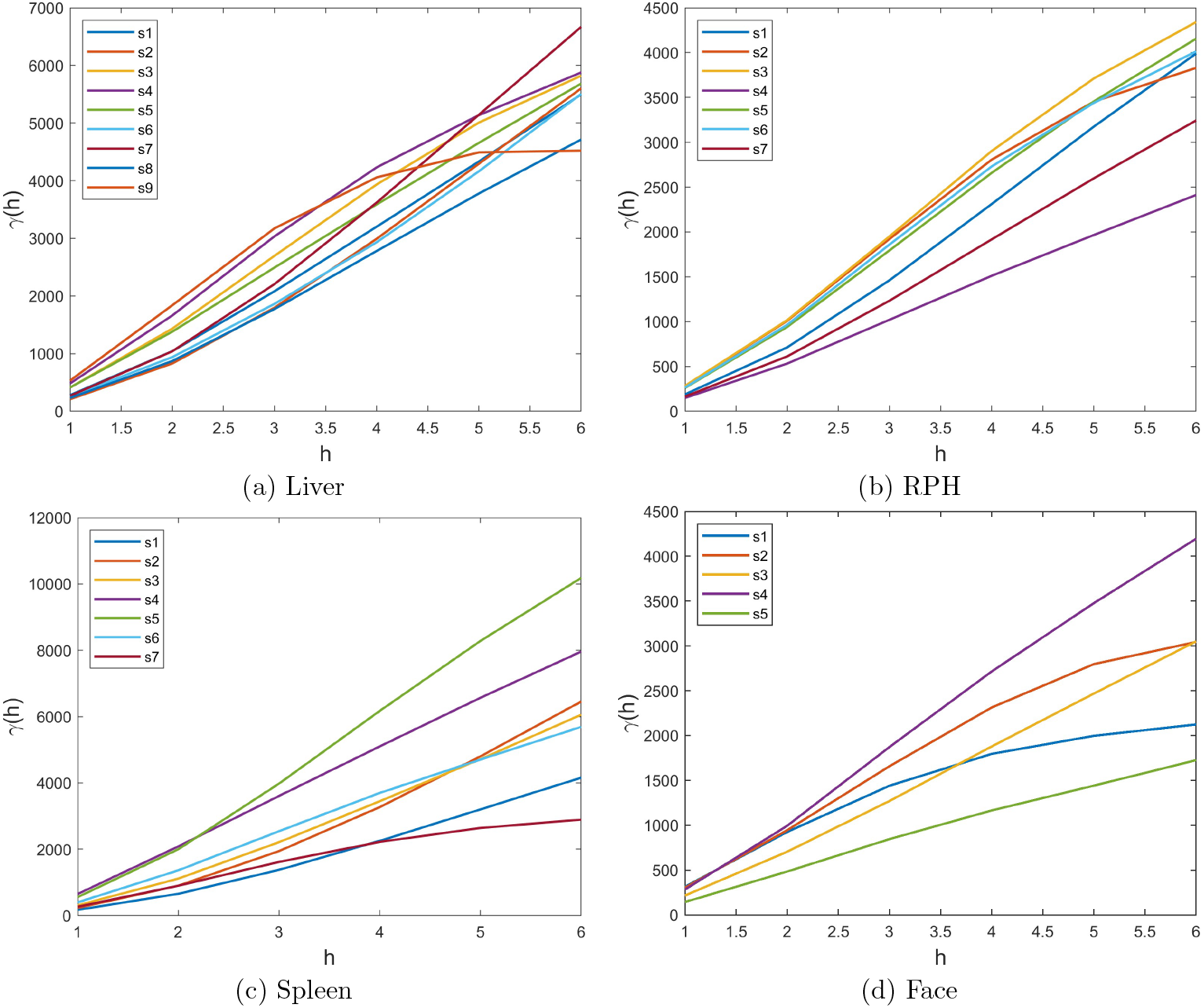
Semi-variograms of extravasation in sequential CT, where the legend indicates CT slice numbers.

Figures 6 and 7 depict the computed FRPs and FRNs derived from the extracted extravasation regions. The FRPs were constructed using an embedding dimension of *m* = 3, a time delay of *τ* = 1, and a clustering parameter set to 12 fuzzy clusters. These parameters were chosen to ensure a robust representation of recurrence structures within the data. The FRNs were subsequently generated using a recurrence threshold of 0.2, ensuring discriminative network connectivities.

**Figure 6:**
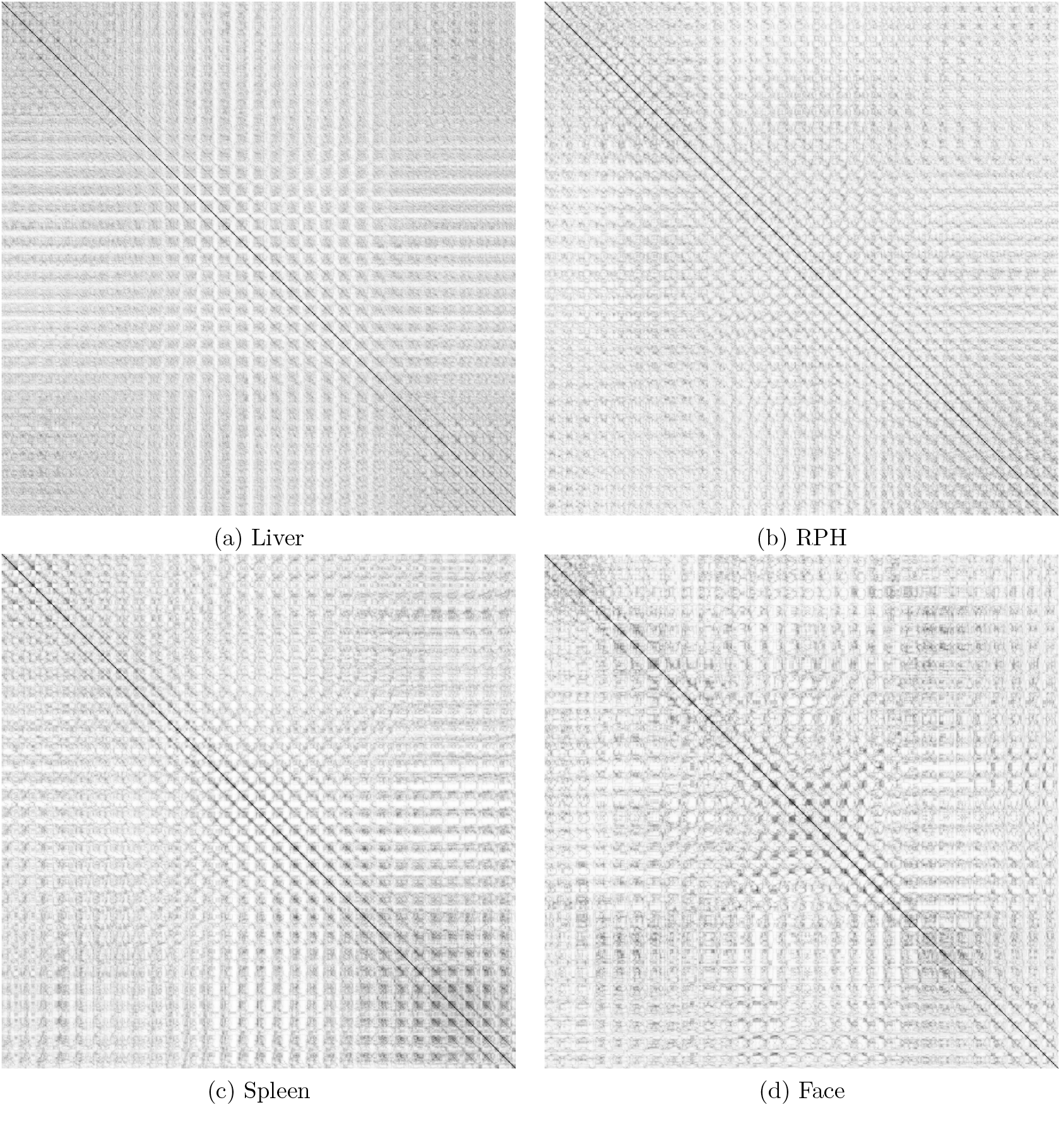
Average FRPs of extravasation in sequential CT.

**Figure 7:**
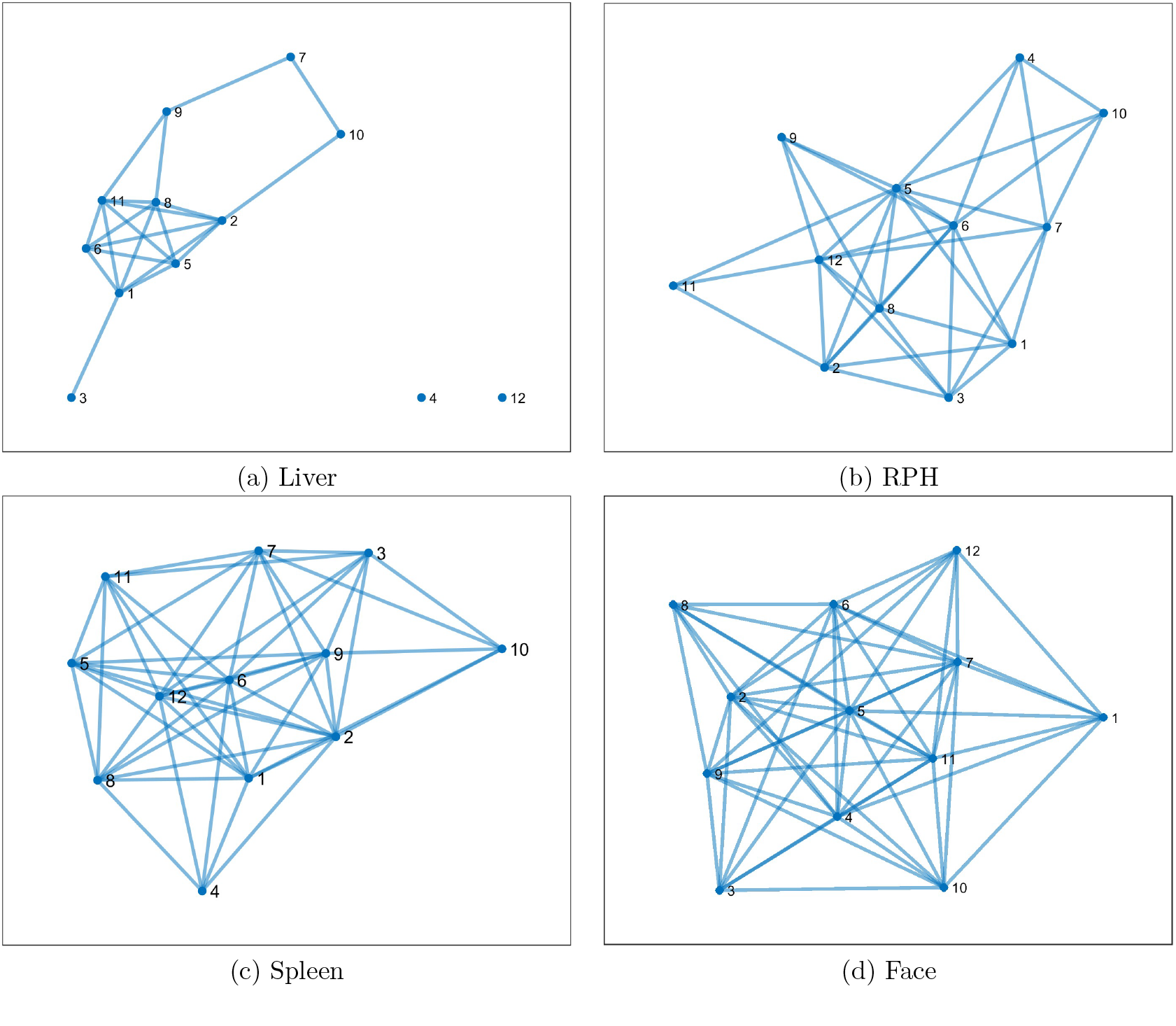
Fuzzy recurrence networks of extravasation in sequential CT.

For the quantification of recurrence dynamics, fuzzy recurrence measures were computed with a minimum diagonal and vertical line length of 5 to maintain statistical relevance. Table 1 summarizes the extracted fuzzy recurrence metrics (fRR, fDET, fLAM, fTT, fDIV, and fENT) for each trauma case, offering a comparative analysis of recurrence-based complexity within different extravasation patterns.

**Table 1:** Fuzzy recurrence measures of extravasation in sequential CT.

| Trauma | $\lambda_{\max}$ | fRR | fDET | fLAM | fTT | fDIV | fENT |
| --- | --- | --- | --- | --- | --- | --- | --- |
| Liver | 6.082 | 1.524e-04 | 0.471 | 0.452 | 3.407 | 0.001 | 1.623 |
| RPH | 5.456 | 5.415e-04 | 0.144 | 0.662 | 2.494 | 0.001 | 1.153 |
| Spleen | 5.779 | 0.002 | 0.464 | 0.673 | 3.970 | 0.002 | 1.760 |
| Face | 6.639 | 0.001 | 0.305 | 0.515 | 2.292 | 0.002 | 1.616 |

Additionally, Table 2 presents two key graph-theoretic properties derived from the FRNs, providing further insights into the topological characteristics of the extravasation structures. These network-based features enable a complementary interpretation of the recurrence dynamics, enhancing the overall understanding of the underlying spatial-temporal patterns in trauma-related contrast extravasation.

**Table 2:** Graph properties of extravasation in sequential CT.

|  | Liver | RPH | Spleen | Face |
| --- | --- | --- | --- | --- |
| Average clustering coefficient | 0.570 | 0.720 | 0.780 | 0.828 |
| Characteristic path length | 1.490 | 1.319 | 1.167 | 1.097 |

## 4 Discussion

The semivariograms of contrast extravasation in liver injury, RPH, spleen injury, and facial trauma reveal distinct spatial variance patterns that reflect differences in contrast dispersion across anatomical regions. In the liver case, the semivariogram exhibits a relatively smooth increase before reaching a well-defined sill (the point at which the spatial variance levels off), suggesting a structured and homogeneous diffusion of contrast medium within the extravasation region. This pattern indicates a consistent spatial correlation of intensity values, likely due to the liver’s highly vascularized tissue and the relatively predictable nature of contrast leakage.

In contrast, the semivariogram for the RPH presents a more gradual rise with a less distinct sill, indicating a broader spatial correlation with diffuse contrast dispersion. The increased variability at larger lag distances suggests heterogeneous bleeding patterns, which may be attributed to the complex anatomical structures within the retroperitoneal space and the presence of multiple tissue compartments that influence contrast movement.

The spleen injury semivariogram displays an intermediate pattern, with a steeper initial rise followed by a moderate leveling off. This suggests a more localized dispersion of contrast with some degree of heterogeneity, potentially influenced by the spleen’s trabecular architecture and its susceptibility to irregular bleeding patterns. Compared to the liver, the semivariogram for the spleen demonstrates a slightly lower spatial correlation range, indicating more localized variations in contrast intensity.

The facial trauma case exhibits the most irregular semivariogram, characterized by an elevated nugget effect and fluctuations throughout the range. This behavior suggests significant microscale heterogeneity and noise, likely due to the intricate vascular network and complex anatomical structures in the face. Unlike the other cases, where contrast dispersion follows a more continuous pattern, the facial trauma semivariogram reflects abrupt intensity variations, possibly due to small-scale vessel ruptures and variable tissue density.

Comparing these semivariograms, it is evident that the liver and spleen injuries demonstrate more structured contrast dispersion, whereas the RPH exhibits a broader, less defined pattern. Facial trauma, in contrast, displays the highest degree of spatial variability, highlighting the complexity of extravasation patterns in this region. These differences underscore the importance of spatial variance analysis in characterizing trauma-related contrast leakage and improving diagnostic interpretation in diverse anatomical contexts.

The FRPs provide a visual representation of contrast extravasation recurrence patterns across different trauma cases, capturing both spatial and temporal complexity. In the liver injury case, the FRP shows an organized pattern with relatively more uniform diagonal structures, indicating higher predictability and temporal coherence in contrast diffusion. This suggests that contrast leakage in the liver follows a consistent pattern, likely influenced by its uniform vascularization.

In contrast, the FRP for the RPH displays a fragmented structure with irregular recurrence points. This fragmentation indicates heterogeneous and unpredictable contrast dispersion, likely due to the complex and compartmentalized retroperitoneal space, where multiple tissue layers and fluid compartments disrupt the continuity of recurrence patterns.

The spleen injury case presents an intermediate recurrence pattern, combining both dense and sparse regions. While the recurrence structure is more stable than that of the RPH, it still shows localized variations in contrast dispersion. The spleen’s trabecular architecture and susceptibility to irregular bleeding possibly contribute to this semi-organized pattern, where contrast leakage maintains some level of coherence but exhibits localized disruptions.

Facial trauma displays the most irregular FRP, characterized by fragmented recurrence structures and a lack of continuous diagonal patterns. This suggests significant spatial and temporal heterogeneity in contrast dispersion, likely due to the intricate vascular network and variable tissue densities in the face. The discontinuous recurrence points reflect the abrupt nature of contrast leakage in facial trauma, where small vessel ruptures lead to unpredictable extravasation dynamics.

Comparing the cases, the liver injury exhibits the most structured recurrence, indicative of stable contrast diffusion. The RPH and spleen injury cases show moderate recurrence organization, reflecting varying levels of spatial heterogeneity. Facial trauma, on the other hand, presents the most chaotic pattern, highlighting the complex and unpredictable nature of extravasation in this region. These findings demonstrate the utility of FRPs in differentiating contrast dispersion behaviors across anatomical sites and provide insights into the underlying mechanisms of trauma-induced vascular injury.

The fuzzy recurrence measures serve as a useful tool for quantifying and characterizing various aspects of the recurrence dynamics of extravasation in the liver injury, RPH, spleen injury, and facial trauma. These measures, which include the *λ*_max_), fRR, fDET, fLAM, fTT, fDIV, and fENT, offer insights into the different patterns and behaviors of recurrence in extravasation associated with respective trauma types, revealing both the underlying complexity of the dynamic systems.

In the case of liver injury, its (*λ*_max_) is the highest among the four trauma types, at 6.082. This high value suggests that the recurrence dynamics of liver injury is relatively more distinct, compared to the other cases. The fRR is quite low at 1.524e-04, indicating sparse recurrence events, which can be interpreted as the presence of less frequent but highly structured recurrence points. The fDET value of 0.471 further emphasizes that while some degree of determinism is present, the system is not completely predictable, reflecting the potential for some irregular behavior in the recurrence pattern. The fLAM value of 0.452 suggests moderate levels of laminarity, indicating that while there is some predictability in the recurrence structure, it is not overwhelmingly regular. The fTT value of 3.407 indicates moderate trapping behavior, suggesting that once recurrence points are visited, they tend to stay within certain regions of the state space for a moderate amount of time before moving to a different area. The very low fDIV value of 0.001 supports the notion that the system’s behavior remains relatively stable, with minimal divergence from the established recurrence patterns. The fENT value of 1.623 is relatively high, indicating a moderate degree of randomness and complexity in the recurrence dynamics of liver injury, despite its overall periodic structure.

For the RPH, the *λ*_max_) is slightly lower at 5.456, signaling a somewhat less distinct recurrence structure compared to the liver injury. The higher fRR of 5.415e-04 suggests that recurrence points occur more frequently, which may indicate a more chaotic recurrence process with less predictability. The fDET value of 0.144, which is the lowest among the cases, further supports this interpretation by indicating a system that behaves more chaotically and is less deterministic. The fLAM value of 0.662 is higher than that of liver injury, suggesting that the recurrence structure of the RPH exhibits more regularity and predictability in certain regions, even if the overall system is less deterministic. The fTT of 2.494 indicates shorter periods of trapping compared to the liver injury, suggesting that recurrence points in the RPH tend to transition more quickly across the state space. The low fDIV value of 0.001 and the relatively low fENT value of 1.153 reflect a less complex system, which is more homogenous in nature. This could be consistent with the disrupted and heterogeneous recurrence patterns seen in hematomas, which are typically characterized by chaotic and irregular dynamics.

In the case of the spleen injury, the *λ*_max_ is intermediate, at 5.779, suggesting a moderate level of recurrence complexity. The fRR of 0.002, higher than both the liver and RPH, suggests more frequent recurrence events, potentially reflecting more regular recurrence patterns that are less chaotic. The fDET value of 0.464 highlights that significant levels of determinism are present, suggesting a system that is relatively predictable, although still somewhat flexible. The fLAM value of 0.673 is the highest among the trauma types, indicating that the spleen injury exhibits a high level of regularity and predictability in its recurrence patterns. The fTT of 3.970 is the highest of all the cases, reflecting pronounced trapping behavior where recurrence points stay within certain areas for a longer period before transitioning. The higher fDIV value of 0.002 and the highest fENT value of 1.760, in comparison to the other trauma types, indicate a system that is highly complex and variable, with both structured and erratic dynamics coexisting in the recurrence process.

Facial trauma, characterized by a complex and irregular recurrence pattern, exhibits the highest *λ*_max_ at 6.639. This value suggests that the recurrence dynamics are highly complex and structured, perhaps reflecting more intricate and multifaceted patterns of extravasation compared to the other trauma cases. The fRR for facial trauma is 0.001, which indicates a relatively sparse recurrence of events, yet the higher complexity in terms of *λ*_max_ suggests that these sparse events are more concentrated and exhibit greater structure. The fDET of 0.305 and fLAM of 0.515 indicate moderate levels of predictability and structure within the recurrence patterns, though not to the extent observed in the spleen injury case. The fTT of 2.292 reflects the relatively short durations that recurrence points tend to stay within certain regions, suggesting a more dynamic and less stable recurrence process compared to the spleen injury. The fDIV value of 0.002 suggests that while the system is somewhat stable, there are still significant deviations from the typical behavior. The fENT of 1.616 further suggests that the recurrence dynamics of facial trauma are marked by a moderate level of randomness, indicating both predictable and erratic features in the recurrence structure.

Having discussed, the fuzzy recurrence measures reveal distinct differences in the dynamics of extravasation across liver injury, retroperitoneal hematoma, spleen injury, and facial trauma. Liver injury shows the most periodic and structured dynamics, with moderate complexity and moderate predictability. RPH, by contrast, exhibits more chaotic behavior, with less determinism and a higher level of randomness. The spleen injury case is characterized by high regularity and predictability, with pronounced trapping behavior, reflecting a highly structured but still complex system. Facial trauma, with the highest *λ*_max_, presents a highly complex and dynamic recurrence structure, marked by moderate levels of predictability but also significant randomness, indicating the intricate and multifaceted nature of extravasation in this trauma type.

The FRNs (Figure 7) presented for four different anatomical regions (liver, RPH, spleen, and facial trauma) exhibit distinct topological characteristics, reflecting underlying structural and dynamical differences in extravasation patterns. By analyzing the average clustering coefficients and characteristic path lengths of the four networks (Table 2), meaningful insights into their connectivity and information flow properties can be inferred.

The average clustering coefficient (ACC) is a measure of how well nodes in a network tend to cluster together, providing insights into the degree of local connectivity. Among the four networks, the facial trauma extravasation network demonstrates the highest ACC (0.828), followed by the spleen (0.780) and RPH (0.720) networks, while the liver injury extravasation network has the lowest ACC (0.570). This suggests that the face network exhibits the most strongly interconnected local structures, forming tightly-knit clusters. The high clustering in the face and spleen networks indicates that their recurrence structures are more locally coherent, potentially reflecting a more predictable pattern of extravasation dynamics in these regions. In contrast, the liver network’s lower clustering suggests a more loosely connected topology, indicating a more dispersed or less predictable recurrence structure. The characteristic path length (CPL) is a measure of the average shortest path between nodes, reflecting the efficiency of information flow across the network. The face network again stands out with the shortest CPL (1.097), suggesting that it has a highly interconnected structure that allows for rapid communication across nodes. The spleen (1.167) and RPH (1.319) networks follow closely, while the liver network has the longest CPL (1.490), implying a less efficient global structure with longer connections between distant nodes.

The observed differences in network topology suggest that extravasation dynamics vary significantly across anatomical regions. The high clustering and short path length of the face and spleen networks indicate a more small-world-like structure [41], which is often associated with a balance between local clustering and global efficiency. These characteristics suggest that extravasation in these regions follows a structured and highly interconnected pattern, potentially due to consistent vascular architecture or localized extravasation phenomena. In contrast, the liver network’s lower clustering and longer path length suggest a more fragmented or less organized recurrence structure. This may indicate that extravasation in the liver is more spatially dispersed, with less predictable local clustering and longer-range connections required to integrate information across the network. The RPH network, with intermediate values for both metrics, may represent a transition between these two extremes, exhibiting moderate clustering while maintaining relatively efficient path lengths.

In summary, this study offers several advantages, including its ability to quantify and characterize the recurrence dynamics of extravasation across various trauma types using spatial statistics and fuzzy recurrence analysis. The approach provides valuable insights into the complex and varied recurrence patterns, revealing distinct differences between liver injury, RPH, spleen injury, and facial trauma. By applying these measures, the study highlights the different levels of determinism, complexity, and predictability in each trauma type, offering a deeper understanding of the underlying mechanisms of extravasation.

However, there are some limitations to consider. The interpretation of fuzzy recurrence measures relies heavily on the quality and resolution of the input data, which in this case, are sequential CT scans. Variations in scan quality or imaging resolution could influence the recurrence dynamics. Additionally, while the study provides a detailed analysis of recurrence behaviors, it does not explore the clinical implications of these findings in terms of treatment or patient outcomes. Another potential drawback is the need for further validation of the fuzzy recurrence measures, especially in clinical settings, to ensure their generalizability and effectiveness across different populations and trauma types.

Future research could focus on refining the fuzzy recurrence methodology to incorporate more diverse and higher-resolution data, such as MRI or PET scans, to enhance the precision of the measures. Furthermore, there is an opportunity to integrate the recurrence analysis with clinical data to better understand the real-world significance of these dynamics in trauma management and recovery. Exploring the relationship between the recurrence measures and patient outcomes, such as recovery time or the risk of complications, would provide valuable insights into how these dynamics might influence clinical decision-making. Lastly, expanding the study to include a larger cohort of trauma cases could help to validate and refine the findings, providing more robust conclusions that could inform future medical interventions.

## 5 Conclusion

This study offers a comprehensive, data-driven analysis of extravasation patterns in various trauma cases, revealing significant spatial and textural characteristics that vary across different organs. These findings reflect the distinct physiological and pathological dynamics of each trauma type, underscoring the complexity and variability inherent in their recurrence behaviors. The study highlights the value of integrating advanced geostatistical and non-linear dynamics methods with CT imaging to provide a deeper, more nuanced understanding of these patterns. By quantifying spatial variability and texture patterns, this approach not only enhances diagnostic precision but also contributes to the early detection of pathological deviations, potentially allowing for more timely and accurate interventions.

The methodology presented here has broad implications for medical imaging, offering valuable insights into spatial analysis that extend beyond the trauma cases examined. As a result, it holds considerable promise for advancing the field of medical diagnostics, particularly in the realm of personalized therapeutic strategies. The ability to identify and analyze complex recurrence dynamics can be applied to various medical conditions, improving the accuracy of early diagnosis and informing tailored treatment plans. Furthermore, this study paves the way for future research to expand on these techniques, incorporating other imaging modalities and clinical data to further refine the analysis and enhance its clinical applicability.

## Author contributions

TDP contributed to the conception of using geostatistics and fuzzy recurrence dynamics for analysis of extravasation on CT, technical design, computer coding and implementation, and writing the original manuscript. MK and TT provided the CT data. All authors contributed to the review, data analysis and interpretation, and approved the final manuscript.

## Ethical approval statement

This study was approved by the Research Ethics Committee of Queen Mary University of London (QME25.0836).

## Data availability

Anonymized CT data are provided on the fist author’s personal homepage: https://sites.google.com/view/tuan-d-pham/codes, under the name “Extravasation in CT”.

## References

1. Pham TD, Tsunoyama T. Exploring extravasation in cancer patients. Cancers (Basel) 2024; 16(13):2308.

2. Roditi G, Khan N, van der Molen AJ, Bellin MF, Bertolotto M, Brismar T, Correas JM, Dekkers IA, Geenen RWF, Heinz-Peer G, Mahnken AH, Quattrocchi CC, Radbruch A, Reimer P, Romanini L, Stacul F, Thomsen HS, Clément O. Intravenous contrast medium extravasation: systematic review and updated ESUR Contrast Media Safety Committee Guidelines. Eur Radiol. 2022; 32(5):3056–3066.

3. Bertoldi K, Bao ACP, Nomura ATG, Glaeser A, de Souza da Silveira JC, Barreto LNM, D’Avila Lauer R, Timponi SCJ. Intravenous contrast media extravasation in patients undergoing computerized tomography scanning in a hospital in southern Brazil: patients profile and possible related causes. Journal of Radiology Nursing 2024; 43(2): 153–157.

4. Moon SN, Pyo JS, Kang WS. Accuracy of contrast extravasation on computed tomography for diagnosing severe pelvic hemorrhage in pelvic trauma patients: a meta-analysis. Medicina (Kaunas) 2021; 57(1):63.

5. Stefanos SS, Kiser TH, MacLaren R, Mueller SW, Reynolds PM. Management of noncytotoxic extravasation injuries: a focused update on medications, treatment strategies, and peripheral administration of vasopressors and hypertonic saline. Pharmacotherapy 2023; 43(4):321–337.

6. Qamar SR, Evans D, Gibney B, Redmond CE, Nasir MU, Wong K, Nicolaou S. Emergent comprehensive imaging of the major trauma patient: a new paradigm for improved clinical decision-making. Can Assoc Radiol J. 2021; 72(2):293–310.

7. Guglielmo FF, Wells ML, Bruining DH, Strate LL, Huete Á, Gupta A, Soto JA, Allen BC, Anderson MA, Brook OR, Gee MS, Grand DJ, Gunn ML, Khandelwal A, Park SH, Ramalingam V, Sokhandon F, Yoo DC, Fidler JL. Gastrointestinal bleeding at CT angiography and CT enterography: imaging atlas and glossary of terms. Radiographics 2021; 41(6):1632–1656.

8. Mericle RA, Lopes DK, Fronckowiak MD, Wakhloo AK, Guterman LR, Hopkins LN. A grading scale to predict outcomes after intra-arterial thrombolysis for stroke complicated by contrast extravasation. Neurosurgery 2000; 46(6):1307–1315.

9. Nagasawa J, Yokoyama T, Fujimoto E, Hozumi M, Kano O. Delayed contrast medium excretion due to renal failure after an emergency mechanical thrombectomy for acute cerebral infarction. Cureus 2024; 16(11) :e74466.

10. Palm H, Lang P, Haentzsch M, Friemert B, Hackenbroch C, Riesner H. Diagnostic accuracy of fluoroscopy, radiography, and computed tomography in detecting cement leakage in kyphoplasty. Journal of J Neurol Surg A Cent Eur Neurosurg 2018; 79(06):502–510.

11. Plat VD, Bootsma BT, Straatman J, van den Bergh J, van Waesberghe JTM, Luttikhold J, Luyer MDP, van der Peet DL, Daams F. The clinical suspicion of a leaking intrathoracic esophagogastric anastomosis: the role of CT imaging. J Thorac Dis. 2020; 12(12):7182–7192.

12. Kim MG, Kim SH, Jeon SK, Han S. Added value of positive intraluminal contrast CT over fluoroscopic examination for detecting gastrointestinal leakage after gastrointestinal surgery. Sci Rep. 2024; 14(1):1011.

13. Yedavalli V, Sammet S. Contrast extravasation versus hemorrhage after thrombectomy in patients with acute stroke. Journal of Neuroimaging 2017; 27(6): 570–576.

14. Juern JS, Milia D, Codner P, Beckman M, Somberg L, Webb T, Weigelt JA. Clinical significance of computed tomography contrast extravasation in blunt trauma patients with a pelvic fracture. J Trauma Acute Care Surg. 2017; 82(1):138–140.

15. Hale O, Deutsch PG, Lahiri A. Epirubicin extravasation: consequences of delayed management. BMJ Case Rep. 2017; 2017:bcr2016218012.

16. Haber ZM, Charles HW, Erinjeri JP, Deipolyi AR. Predictors of Active Extravasation and Complications after Conventional Angiography for Acute Intraabdominal Bleeding. J Clin Med. 2017; 6(4):47.

17. Ding S, Meystre NR, Campeanu C, Gullo G. Contrast media extravasations in patients undergoing computerized tomography scanning: a systematic review and meta-analysis of risk factors and interventions. JBI Database System Rev Implement Rep. 2018; 16(1):87–116.

18. Ye Z, Ai X, Zheng J, Hu X, You C, Andrew M F, Fang F. Extravasation of contrast (Spot Sign) predicts in-hospital mortality in ruptured arteriovenous malformation. Br J Neurosurg. 2019; 33(2):149–155.

19. Dreizin D, Liang Y, Dent J, Akhter N, Mascarenhas D, Scalea TM. Diagnostic value of CT contrast extravasation for major arterial injury after pelvic fracture: a meta-analysis. Am J Emerg Med. 2020; 38(11):2335–2342.

20. Hirata I, Mazzotta A, Makvandi P, Cesini I, Brioschi C, Ferraris A, Mattoli V. Sensing technologies for extravasation detection: a review. ACS Sensors 2023; 8 (3): 1017–1032.

21. Liu W, Wang P, Zhu H, Tang H, Guan H, Wang X, Wang C, Qiu Y, He L. Contrast media extravasation injury: a prospective observational cohort study. Eur J Med Res. 2023; 28(1):458.

22. Mahajan A, Gupta A, Shukla S, Agarwal U, Rai P, Sable N, Venugopal AP, Sunthar P, Banwar P, Rane PB, Thakur M. Additional use of extrinsic warmer for intravenous CT contrast media and its impact on incidence of contrast extravasations and allergic like reactions: a prospective observational case control study. Clin Radiol. 2024; 79(11):851–860.

23. Wang L, Chen Q, Liu H, Wang X, Qian Q, Xu M, Ma L, Wang X. Frequency and risk factors of contrast media extravasation in 378,082 intravenous contrast-enhanced CT scans. Eur J Radiol. 2025; 184:111992.

24. Kobayashi N, Nakaura T, Shiraishi K, Uetani H, Nagayama Y, Kidoh M, Oda S, Sakabe D, Ikeda R, Hatemura M, Murakami M, Funama Y, Hirai T. A novel approach to detecting contrast extravasation in computed tomography: evaluating the injection pressure-to-injection rate ratio. J Comput Assist Tomogr. 2025; 49(1):125–132.

25. J.P. Chiles, P. Delfiner. Geostatistics: Modeling Spatial Uncertainty, second edition. John Wiley& Sons Hoboken: New Jersey, 2012.

26. Pham TD. Fuzzy recurrence plots. EPL 2016; 116:50008.

27. Pham TD. Fuzzy Recurrence Plots and Networks with Applications in Biomedicine. Springer Nature: Switzerland, 2020.

28. Pham TD, Holmes SB, Patel M, Coulthard P. Features and networks of the mandible on computed tomography. R. Soc. Open Sci. 2024; 11:231166.

29. Pham TD, Kitamura M, Tsunoyama T. Recurrence dynamics of extravasation on computed tomography. 2025 IEEE International Conference on Cybernetics and Innovations (ICCI 2025), 6 pages.

30. Krige DG. A statistical approach to some basic mine valuation problems on the Witwatersrand. J. of the Chem., Metal. and Mining Soc. of South Africa 1951; 52 (6):119–139.

31. Matheron G. Estimating and Choosing, Springer-Verlag: Berlin, 1989.

32. Isaaks EH, Srivastava RM. (1989), An Introduction to Applied Geostatistics. Oxford University Press: New York, 1989.

33. Bezdek JC. Pattern Recognition with Fuzzy Objective Function Algorithms. Plenum Press, New York: NY, 1981.

34. Zadeh LA. Similarity relations of fuzzy orderings. Inform. Sci. 1971; 3:177–200.

35. Pham TD. Quantification analysis of fuzzy recurrence plots. EPL 2022; 137: 62002.

36. Zbilut JP, Webber CL. Embeddings and delays as derived from quantification of recurrence plots. Physics Letters A 1992; 171:199–203.

37. Webber CL, Zbilut JP. Dynamical assessment of physiological systems and states using recurrence plot strategies. J Appl Physiol. 1994; 76:965–973.

38. Marwan N, Wessel N, Meyerfeldt U, Schirdewan A, Kurths J. Recurrence-plot-based measures of complexity and their application to heart-rate-variability data. Phys Rev E Stat Nonlin Soft Matter Phys. 2002; 66(2 Pt 2):026702.

39. Marwan N, Kurths J. Line structures in recurrence plots. Phys. Lett. A 2005; 336: 349–357.

40. Pham TD. From fuzzy recurrence plots to scalable recurrence networks of time series. EPL 2017; 118: 20003.

41. Watts DJ, Strogatz S. Collective dynamics of “small-world” networks. Nature 1998; 393:440–442.

42. Albert R, Barabasi AL. Statistical mechanics of complex networks. Rev. Mod. Phys. 2002; 74:47–97.

43. Pham TD. Convolutional fuzzy recurrence eigenvalues. EPL 2021; 135: 20002.

